# EFFECT OF CONVALESCENT PLASMA ON MORTALITY IN PATIENTS WITH COVID-19 PNEUMONIA

**DOI:** 10.1101/2020.10.08.20202606

**Authors:** Martín R. Salazar, Soledad E. González, Lorena Regairaz, Noelia S. Ferrando, Veronica V. González Martínez, Patricia M. Carrera Ramos, Laura Muñoz, Santiago A. Pesci, Juan M. Vidal, Nicolas Kreplak, Elisa Estenssoro

## Abstract

**Background:** Convalescent plasma, widely utilized in viral infections that induce neutralizing antibodies, has been proposed for COVID-19, and preliminary evidence shows that it might have beneficial effect. Our objective was to compare epidemiological characteristics and outcomes between patients who received convalescent plasma for COVID-19 and those who did not, admitted to hospitals in Buenos Aires Province, Argentina, throughout the pandemic.

**Methods:** This is a multicenter, retrospective cohort study of 2-month duration beginning on June 1, 2020, including unselected, consecutive adult patients with diagnosed COVID-19, admitted to 215 hospitals with pneumonia. Epidemiological and clinical variables were registered in the Provincial Hospital Bed Management System. Convalescent plasma was supplied as part of a centralized, expanded access program.

**Results:** We analyzed 3,529 patients with pneumonia, predominantly male, aged 62±17, with arterial hypertension and diabetes as main comorbidities; 51.4% were admitted to the ward, 27.1% to the Intensive Care Unit (ICU), and 21.7% to the ICU with mechanical ventilation requirement (ICU-MV). 28-day mortality was 34.9%; and was 26.3%, 30.1% and 61.4% for ward, ICU and ICU-MV patients. Convalescent plasma was administered to 868 patients (24.6%); their 28-day mortality was significantly lower (25.5% vs. 38.0%, p<0.001). No major adverse effects occurred.

Logistic regression analysis identified age, ICU admission with and without MV requirement, diabetes and preexistent cardiovascular disease as independent predictors of 28-day mortality, whereas convalescent plasma administration acted as a protective factor.

**Conclusions:** Our study suggests that the administration of convalescent plasma in COVID-19 pneumonia admitted to the hospital might be associated with decreased mortality.

**Key Points:**

- Preliminary evidence showed that convalescent plasma might be beneficial in COVID-19.
- In a cohort of 3,529 patients with pneumonia due to COVID-19, convalescent plasma was administered to 868 patients, without major adverse effects.
- Convalescent plasma was independently associated with decreased mortality.

## Introduction

In December 2019 in Wuhan, China, the first cases of pneumonia caused by SARS-CoV-2, a novel coronavirus, were reported; the disease was subsequently named COVID-19. The new virus spread across the world relentlessly, and on March 11 the World Health Organization declared COVID-19 a pandemic. Up to now, COVID-19 cases are approaching 30,000,000 with 935,000 dead [1,2].

Few treatments have proven effective for COVID-19 [3]. The administration of convalescent plasma, widely utilized in viral infections that induce neutralizing antibodies, has also been proposed [4-6]. It was used during outbreaks of severe acute respiratory disease caused by other coronaviruses, SARS-CoV-1 and MERS-CoV, with varying results and when administered early, it decreased length of hospital stay [7-9]. Convalescent plasma utilization has an acceptable safety profile and its administration constitutes a feasible approach to implement during a pandemic, even in low-resource settings. In COVID-19, it might reduce viral burden, improve clinical status, and decrease mortality [10-12]. On March 24, 2020, the Food and Drug Administration of the United States launched an Expanded Access Program to collect convalescent plasma donated by individuals who had recovered from COVID-19, and on August 23 approved emergency use [13]. A study conducted in 20,000 patients confirmed the safety of convalescent plasma and, thereafter, in a study of 30,000 patients, the same group of researchers demonstrated a decrease in mortality when convalescent plasma was administered early in the course of COVID-19 [14-15].Convalescent plasma is currently being evaluated in 126 clinical trials [16].

Early in the emergency caused by the COVID-19 pandemic, the Ministry of Health of the Province of Buenos Aires, Argentina, created the Centralized Registry of Convalescent Plasma Donors (CROCPD-BA), with the aim of collecting, processing and distributing convalescent plasma, and issuing recommendations for its use in patients with COVID-19 [17]. Accordingly, the objective of the present study is to compare the epidemiological characteristics, outcomes and independent predictors of mortality among patients who received convalescent plasma and those who did not receive it, who were admitted to hospitals in Buenos Aires Province for COVID-19 throughout the pandemic.

## Methods

This was a multicenter retrospective cohort study conducted over 2 months, beginning on June 1, 2020, which included consecutive patients ≥18 years diagnosed with SARS CoV-2 with RT-PCR, admitted to hospitals with pneumonia. Data were obtained from the National Vigilance System (SNVS 2.0), the Provincial Hospital Bed Management System, and the CROCPD-BA.

Collected variables were age, gender, comorbidities [18-19] (arterial hypertension, diabetes, preexistent cardiovascular disease, chronic obstructive pulmonary disease, immunodeficiency), requirement of mechanical ventilation, treatments, death or discharge, and convalescent plasma administration. Severe adverse events related to plasma infusion, as transfusion-related acute lung injury (TRALI) and transfusion-associated circulatory overload (TACO) were also recorded [20].

Information about plasma collection and characteristics is available in the Supplement. The requirement of convalescent plasma was initiated by assistant physicians as part of a Program of Expanded Access [17]. The indications issued by the CROCPD-BA were presence of pneumonia, defined as of lung infiltrates, plus one of the following:

- Dyspnea with respiratory rate ≥ 30 breaths/minute
- Oxygen saturation ≤93%
- Oxygen requirement
- PaO_2_FIO_2_ <300 mmHg
- Increase in lung infiltrates >50% during the previous 24-48 hours
- Alteration in consciousness
- Multiple organ dysfunction
- Age >65 years
- Any of the above mentioned comorbidities

All units of transfused convalescent plasma had an Ig-G antibody titer ≥1:400. The infused volume per unit was 200-250 ml.

Initial severity of illness was assessed according to the site of admission: general ward, Intensive Care Unit (ICU), and ICU admission with requirement of mechanical ventilation (ICU-VM). The main outcome variable was 28-day mortality. Deaths due to COVID-19 were confirmed on patient death certificates.

### Statistical analysis

Continuous variables were expressed as mean ± standard deviation (SD) or median, [0.25-0.75] percentiles. Categorical variables were expressed as percentages. Differences between survivors and nonsurvivors, and between patients who received plasma or not, were analyzed with chi-square, t, or Mann-Whitney U-tests, as appropriate.

To identify independent predictors of 28-day mortality, variables differing between survivors and nonsurvivors with a *p* value <0.10 were entered into a multivariable regression model, using a forward stepwise analysis. Adjusted risks were expressed as odd ratios (OR) and confidence intervals of 95% [CI95%]

Data were analyzed with SSPS-21 (Amonk, NY, US).

A two-tailed p value <0.05 was considered significant.

This study was approved by the Central Ethics Committee of the Ministry of Health of Buenos Aires Province (Expedient 2020-14965594). The resolution 103/2017 of the Ministry of Health of the Province of Buenos Aires establishes the obligation of registration and accreditation of all the Institutional Ethics Committees at the Central Ethics Committee of the Ministry of Health the Province of Buenos Aires; which is not associated with any institution or organization except the same Ministry, as it is the Ethics Committee of the said body, and evaluates all projects developed by institutions of the Ministry.

In the protocol of the present study, the Central Ethics Committee acts as an Institutional Evaluation Committee in use of the powers provided for by Decree 3385/08 as a research project, in which the Ministry of Health of the Province of Buenos Aires acts both as sponsor and center.

The Central Committee established that this observational study had an adequate risk-benefit ratio and requested the anonymization of data.

The administration of convalescent plasma required signed consent from each patient or legal representative, according to CROCPD-BA regulations (Expedient 2919/2123/2020).

## Results

During the study period, 3,529 patients with COVID-19 pneumonia were admitted to 215 hospitals. Epidemiological data of the entire group and comparisons between survivors and nonsurvivors are shown in Table 1. Briefly, this was a predominantly male population, aged 62±17, with arterial hypertension and diabetes as main comorbidities. With respect to disease severity, 51.4% were admitted to the ward, 27.1% to the ICU without mechanical ventilation need, and 21.7% to the ICU, with mechanical ventilation requirement (ICU-MV).

**Table 1.**
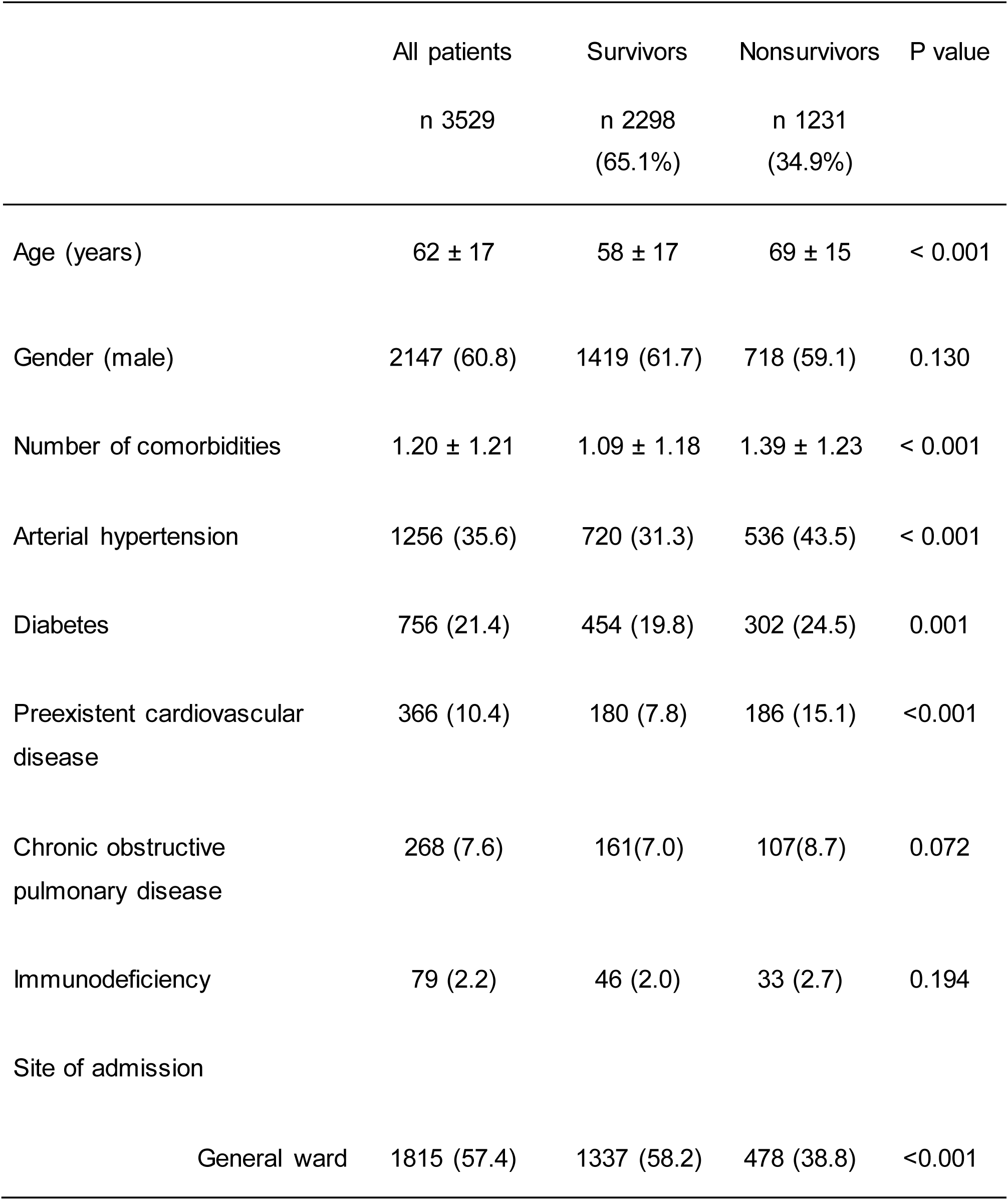
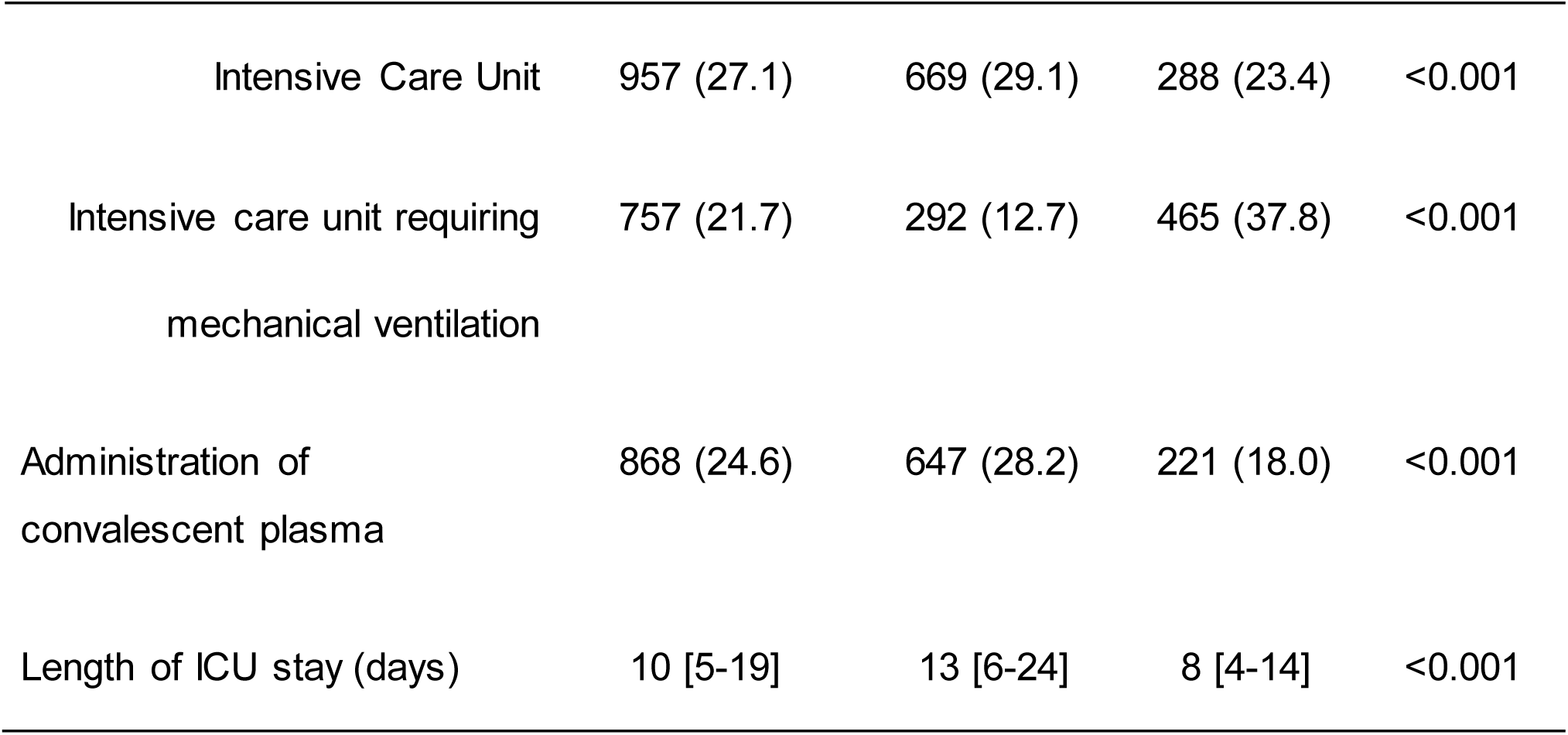
Characteristics of the entire group, and comparison between survivors and nonsurvivors.

|  | All patients<br>n 3529 | Survivors<br>n 2298<br>(65.1%) | Nonsurvivors<br>n 1231<br>(34.9%) | P value |
| --- | --- | --- | --- | --- |
| Age (years) | 62 ± 17 | 58 ± 17 | 69 ± 15 | < 0.001 |
| Gender (male) | 2147 (60.8) | 1419 (61.7) | 718 (59.1) | 0.130 |
| Number of comorbidities | 1.20 ± 1.21 | 1.09 ± 1.18 | 1.39 ± 1.23 | < 0.001 |
| Arterial hypertension | 1256 (35.6) | 720 (31.3) | 536 (43.5) | < 0.001 |
| Diabetes | 756 (21.4) | 454 (19.8) | 302 (24.5) | 0.001 |
| Preexistent cardiovascular disease | 366 (10.4) | 180 (7.8) | 186 (15.1) | <0.001 |
| Chronic obstructive pulmonary disease | 268 (7.6) | 161(7.0) | 107(8.7) | 0.072 |
| Immunodeficiency | 79 (2.2) | 46 (2.0) | 33 (2.7) | 0.194 |
| Site of admission |  |  |  |  |
| General ward | 1815 (57.4) | 1337 (58.2) | 478 (38.8) | <0.001 |
| Intensive Care Unit | 957 (27.1) | 669 (29.1) | 288 (23.4) | <0.001 |
| Intensive care unit requiring<br>mechanical ventilation | 757 (21.7) | 292 (12.7) | 465 (37.8) | <0.001 |
| Administration of<br>convalescent plasma | 868 (24.6) | 647 (28.2) | 221 (18.0) | <0.001 |
| Length of ICU stay (days) | 10 [5-19] | 13 [6-24] | 8 [4-14] | <0.001 |

Twenty-eight-day mortality was 34.9% for the entire group; and respectively, for ward, ICU and ICU-MV patients was 26.3%, 30.1% and 61.4%. Survivors were significantly younger, had less comorbidities, lower admission to the ICU, and had received plasma more frequently.

Convalescent plasma was administered to 868 patients (24.6%) (Table 2). Compared to the remaining 2,298, this group was composed of younger and predominantly male patients, with higher prevalence of arterial hypertension, diabetes, and higher ICU admission. The rate of mechanical ventilation use was similar in both groups.

**Table 2.** Characteristics of patients receiving and non-receiving convalescent plasma.

|  | No Plasma<br>n 2298 (65.1%) | Plasma<br>n 868 (34.9) | P value |
| --- | --- | --- | --- |
| Age (years) | 64 ± 17 | 56 ± 13 | < 0.001 |
| Gender (male) | 1547 (58.1) | 600 (69.1) | < 0.001 |
| Number of comorbidities | 1.11 ± 1.23 | 1.55 ± 1.06 | < 0.001 |
| Arterial hypertension | 914 (34.3) | 342 (39.4) | 0.007 |
| Diabetes | 532 (20.0) | 224 (25.8) | < 0.001 |
| Preexistent cardiovascular disease | 282 (10.6) | 84 (9.7) | 0.440 |
| Chronic obstructive pulmonary disease | 201 (8.7) | 67 (7.7) | 0.873 |
| Immunodeficiency | 59 (2.2) | 20 (2.3) | 0.880 |
| Site of admission |  |  |  |
| General ward | 1409 (53) | 406 (46.8) | 0.002 |
| Intensive Care Unit | 677 (25.4) | 280 (32.3) | < 0.001 |
| Intensive care unit requiring<br>mechanical ventilation | 575 (21.6) | 182 (21.0) | 0.690 |
| 28-day mortality | 1010 (38.0) | 221 (25.5) | < 0.001 |
| Length of ICU stay (days) | 10 [4-17] | 12 [7-18] | <0.001 |

Twenty-eight-day unadjusted mortality was significantly lower in the entire group of patients receiving convalescent plasma, compared to those who had not (25.5% vs. 38.0%; OR 0.59 [0.47-0.66], p<0.001); and also in patients in the ward (14.0% vs. 29.9%; OR 0.38 [0.28-0.52], p<0.001), and in the ICU-MV (50.0% vs. 65.0%; OR 0.54 [0.38-0.75], p<0.001). Patients in the ICU without-MV had a trend to decreased mortality (26.1% vs. 31.8%; OR 0.76 [0.56-1.03], p=0.081).

Logistic regression analysis identified age, ICU admission with and without MV, diabetes and preexistent cardiovascular disease as independent predictors of 28-day mortality, while convalescent plasma administration was associated with decreased mortality (Table 3).

**Table 3.** Independent predictors of 28-day mortality, as identifies with logistic regression analysis.

|  | OR | 95% C.I. | P value |
| --- | --- | --- | --- |
| Admission to the ICU<br>(vs. admission to the ward) | 1.298 | 1.079 - 1.563 | 0.006 |
| Admission to the ICU with MV requirement<br>(vs. admission to the ward) | 5.907 | 4.848 – 7.199 | < 0.001 |
| Preexistent cardiovascular disease | 1.452 | 1.141 - 1.848 | 0.002 |
| Diabetes | 1.320 | 1.098 - 1.587 | 0.003 |
| Age (per year) | 1.046 | 1.040 - 1.052 | < 0.001 |
| Administration of convalescent plasma | 0.756 | 0.622 - 0.920 | 0.005 |

Abbreviations. ICU (Intensive Care Unit); MV (mechanical ventilation)

## Discussion

The main finding of our study was that the administration of convalescent plasma to patients with COVID-19 pneumonia was associated with a decrease of 24.4% in adjusted mortality. This effect was consistent over all grades of severity on admission, although it was greater in less critical patients—those admitted to the general ward.

In this study, the global mortality of 34.6% was higher than the 21-28% shown in observational studies [21-24] which can be ascribed to a different patient case-mix. The proportion of patients admitted to the ICU was 42.6%, of which 21.6% required mechanical ventilation on admission. These figures are notably higher than those reported by two studies from Spain (respectively for each: n=15,111 and 4,035, with ICU admission of 8.3% and 18%;and mortality of 21% and 28%); United States (n=11,721, ICU admission of 19.9%, and mortality of 21.4%), and United Kingdom (n= 20,133, ICU admission 16.8%, and mortality of 26%) [21-24].

The efficacy of convalescent plasma in COVID-19 has been subject to much debate, due to the lack of a clinical trial with sufficient power to confirm it. For example, a study carried out in Wuhan was prematurely terminated due to the end of the pandemic, although significant clinical improvement was observed in patients with severe disease [10]. Likewise, a study from The Netherlands was stopped because 79% of patients already had high titers of neutralizing antibodies before receiving convalescent plasma [25]. A recent clinical trial from India which excluded critically ill patients did not find any clinical benefit. However, these results might be ascribed to the absence of neutralizing antibodies or to titers lower than 1:80 in 27% and 45% of convalescent plasma units, respectively [26]. Moreover, 86% of patients in the plasma subgroup had detectable neutralizing antibodies on enrollment; so it is uncertain if the intervention would have been efficacious.

Conversely, two small clinical trials demonstrated a significant decrease in mortality: in a study from Spain (n=81) including severely ill patients, mortality in the convalescent plasma subgroup was 0% vs. 9.3 % in the control, and in an Iraqi study (n =49), it was 4.8% vs. 28.5%, respectively [27-28].

Many observational studies support a probable efficacy of convalescent plasma. For example, a case-control study from China (including 138 cases and 1,568 controls) reported 2.2% mortality for the convalescent plasma subgroup, versus 4.1% for the control [29]. Furthermore, in a case-control study from the US including non-ventilated patients, 14-day mortality was 12.8% in the subgroup that had received convalescent plasma, vs. 24.4% in the control [30]. Similar results were reported in a matched case-control study, also from the US (136 cases, 251 controls), which showed lower mortality in patients receiving early administration of convalescent plasma with high titers of antibodies: 1.2% vs 8.9% [31]. Finally, the large case-series from the Mayo Clinic (n=35,322) showed a relative risk of 30-day mortality of 0.77 [0.63-0.94] among patients transfused with plasma units of high antibody titers, compared to those transfused with low titers [15].

Our study develops a different approach to this very relevant issue. We analyzed a cohort of 3,529 unselected, consecutive patients with COVID-19 pneumonia, of whom 868 received convalescent plasma; its administration was evaluated as any other prognostic variable for mortality. We observed an independent, favorable effect on survival, and this is a novel finding. Although the nature of our study was observational, it was carried out using a robust database composed of observations prospectively collected, within the framework of a pre-established government program. Other independent predictors of mortality were age, diabetes and cardiovascular disease, similar to current literature on the topic [23,32-34].

This effect of convalescent plasma was more pronounced in less severe patients—those admitted to the ward, suggesting the importance of timely administration. Even though age >65 was one inclusion criterion for receiving convalescent plasma, surprisingly, those who received it were, in fact, younger. We cannot discard selection bias of physicians prescribing a seemingly promising therapy to patients with greater chances of responding to it. Nevertheless, older age was an independent predictor of mortality, as expected [23,34]

### Limitations

The main limitation of this study is the lack of randomized assignment of convalescent plasma administration. Additionally, unmeasured confounders might have influenced the results, such as other risk factors or treatments. Since severity of illness on admission could not be evaluated with an established score, misclassification of patients might have occurred. However, the use of severity of illness on admission as a surrogate of acuity has already been utilized [3]. A more detailed analysis of the clinical variables collected could not be done, because of the type of data recorded in the register. Finally, the reason why assistant physicians chose not to administer convalescent plasma to patients with COVID-19 pneumonia fulfilling the inclusion criteria are unknown, but we speculate that some physicians might have felt uncomfortable with prescribing an experimental treatment to their patients.

In conclusion, our study suggests that the administration of convalescent plasma in COVID-19 pneumonia might be associated with decreased mortality. Large, well-designed clinical trials are required to confirm these findings.

## Supporting information

https://www.dropbox.com/s/0lx36zp4ybo7a3z/Supplementary%20Data.%20EFFECT%20OF%20CONVALESCENT%20PLASMA%20ON%20MORTALITY%20IN%20PATIENTS%20WITH%20COVID-

## Data Availability

I declare that all the data referred to in the manuscript are available.

## NOTES

### Authors’ contributions

M.R.S., E.E. contributed study design, analysis, interpretation, and writing. Supervision or mentorship. S.E.G, L.R performed literature search, study design, and writing.N.F, L.M, P.M.C.R, S.A.P, J.M.V. contributed data collection and analysis. V.M.G carried out scientific management of the project. N.K. research idea and study design

## Acknowledgements

Luis Cantaluppi, Yanina Spinelli, Cecilia Girard Bosch, Patricia Méndez, Andrea Gamarnik, Gian Pietro Fernández Rojas, Alejandra Debonis, Laura Vives, Verónica Copolillo, Vanesa Fernandez, Natalia Nuñez, Ariel Sola, Alejandra Toledo, Miriam Cardalda, Agustina Lagrava, Rosario Céspedes, Jorgelina Aberer, Silvana Gabriela Turano, Carlos Antonio Cuchetti, Agustina Martins, Luciana Spizzirri, Gustavo Mariño, Andrea Lopardi, Cecilia Avatte, Viviana Falasco, Martin Fabian Ruiz, Ana Laura Gonzalez, Gabriela Rodriguez, Horacio Rubén López Alegre, Pablo Schon, Mariana Casalins, Gustavo Cañete, Soledad Vallejo, Karina Sarmiento, Christian Martinez Kovach, Fabiana Ferrero, Liliana Disalvo, Ana Varea, Ignacio Méndez, Lucrecia Fotia, Marcela Padua, Isabel Ballesteros, Adriana Hilda Romani, Daniela Federico, Diego Ortale, Viviana Jalife, Josefina Viaño, Mariana Jaime, Federico Gorini, Sandra González, Analia Vairo, Marina Alvarez, Pablo Marocco, Diego Saez, Federico Paicher y Marcelo Cordero.

The authors would especially like to acknowledge the collaboration of Mario Rovere (School of Health Government Affairs), Francisco Leone (Unique Implant Ablation Coordinator Center of Buenos Aires) and Juan Sebastián Riera (Provincial Direction of Hospitals), without them this work would not have been possible.

## References

1. WHO. Cronolog a de la respuesta de la OM a la COVID-19. 2020. Available at:https://www.who.int/es/news-room/detail/29-06-2020-covidtimeline. Accessed 25 September 2020.

2. COVID-19 Map - Johns Hopkins Coronavirus Resource Center. 2020. Available at:https://coronavirus.jhu.edu/map.html. Accessed 25 September 2020.

3. RECOVERY Collaborative Group, et al. Dexamethasone in Hospitalized Patients with Covid-19 - Preliminary Report [published online ahead of print, 2020 Jul 17]. N Engl J Med. 2020;NEJMoa2021436. doi:10.1056/NEJMoa2021436

4. Marano G, Vaglio S, Pupella S, et al. Convalescent plasma: new evidence for an old therapeutic tool? Blood Transfus. 2016 Mar;14(2):152–7. doi: 10.2450/2015.0131-15.

5. Wooding DJ, Bach H. Treatment of COVID-19 with convalescent plasma: lessons from past coronavirus outbreaks. Clin Microbiol Infect. 2020 Oct;26(10):1436–1446. doi: 10.1016/j.cmi.2020.08.005.

6. Psaltopoulou T, Sergentanis TN, Pappa V, et al. The Emerging Role of Convalescent Plasma in the Treatment of COVID-19. HemaSphere. 2020 May 21;4(3):e409. doi:10.1097/HS9.0000000000000409.

7. Cheng Y, Wong R, Soo YO, et al. Use of convalescent plasma therapy in SARS patients in Hong Kong. Eur J Clin Microbiol Infect Dis. 2005 Jan;24(1):44–6. doi: 10.1007/s10096-004-1271-9.

8. Ko JH, Seok H, Cho SY, et al. Challenges of convalescent plasma infusion therapy in Middle East respiratory coronavirus infection: a single centre experience. Antivir Ther. 2018;23(7):617–622. doi:10.3851/IMP3243

9. Yeh KM, Chiueh TS, Siu LK, et al. Experience of using convalescent plasma for severe acute respiratory syndrome among healthcare workers in a Taiwan hospital. J Antimicrob Chemother. 2005 Nov;56(5):919–22. doi: 10.1093/jac/dki346.

10. Li L, Zhang W, Hu Y, et al. Effect of Convalescent Plasma Therapy on Time to Clinical Improvement in Patients With Severe and Life-threatening COVID-19: A Randomized Clinical Trial. JAMA. 2020 Aug 4;324(5):460–470. doi:10.1001/jama.2020.10044

11. Joyner M, Klassen S, Senefeld J, et al. Evidence favouring the efficacy of convalescent plasma for COVID-19 therapy. medRxiv [Preprint] Aug 28, 2020 [cited 2020 Sep 25]. Available from: doi: 10.1101/2020.07.29.20162917

12. Salazar E, Christensen PA, Graviss EA, et al. Treatment of Coronavirus Disease 2019 Patients with Convalescent Plasma Reveals a Signal of Significantly Decreased Mortality. Am J Pathol. 2020 Aug 11:S0002-9440(20)30370-9. doi: 10.1016/j.ajpath.2020.08.001. Epub ahead of print.

13. FDA Issues Emergency Use Authorization for Convalescent Plasma as Potential Promising COVID–19 Treatment, nother chievement in dministration’s Fight Against Pandemic. 2020. Available at:https://www.fda.gov/news-events/press-announcements/fda-issues-emergency-use-authorization-convalescent-plasma-potential-promising-covid-19-treatment. Accessed 25 September 2020.

14. Joyner MJ, Bruno KA, Klassen SA, et al. Safety Update: COVID-19 Convalescent Plasma in 20,000 Hospitalized Patients. Mayo Clin Proc. 2020 Sep;95(9):1888–1897. doi: 10.1016/j.mayocp.2020.06.028.

15. Joyner MJ, Senefeld JW, Klassen SA, et al. Effect of Convalescent Plasma on Mortality among Hospitalized Patients with COVID-19: Initial Three-Month Experience. medRxiv [Preprint]. Aug 12, 2020 [cited 2020 Sep 25]. doi: 10.1101/2020.08.12.20169359.

16. Home - ClinicalTrials.gov. 2020. Available at:https://clinicaltrials.gov/ct2/home. Accessed 16 September 2020.

17. Gobierno de la Provincia de Buenos Aires. Emergencia Sanitaria. Registro Único de Donantes de Plasma Convaleciente de la Provincia de Buenos Aires (RUDPCBA) para la obtención, procesamiento, distribución y recomendaciones terapéuticas sobre su uso en el tratamiento de pacientes con COVID-19.Available at: https://portal-coronavirus.gba.gob.ar/es/efectores-de-salud. Accesed September 16 2020

18. Guan W, Ni Z, Hu Y, Liang W, et al. Clinical Characteristics of Coronavirus Disease 2019 in China. N Engl J Med. 2020 Feb 28 : NEJMoa2002032. Published online 2020 Feb 28. doi: 10.1056/NEJMoa2002032

19. Grasselli G, Zangrillo A, Zanella A et al. Baseline Characteristics and Outcomes of 1591 Patients Infected With SARS-CoV-2 Admitted to ICUs of the Lombardy Region, Italy JAMA. 2020 Apr 28; 323(16): 1574–1581

20. Narick C, Triulzi DJ, Yazer MH. Transfusion-associated circulatory overload after plasma transfusion. Transfusion. 2012 Jan;52(1):160–5. doi: 10.1111/j.1537-2995.2011.03247.

21. Casas-Rojo JM, Antón-Santos JM, Millán-Núñez-Cortés J, et al. Clinical characteristics of patients hospitalized with COVID-19 in Spain: Results from the SEMI-COVID-19 Registry. Rev Clin Esp. 2020:S0014-2565(20)30206-X. English, Spanish. doi: 10.1016/j.rce.2020.07.003. Epub ahead of print.

22. Fried MW, Crawford JM, Mospan AR, et al. Patient Characteristics and Outcomes of 11,721 Patients with COVID19 Hospitalized Across the United States. Clin Infect Dis. 2020:ciaa1268. doi: 10.1093/cid/ciaa1268. Epub ahead of print.

23. Docherty Harrison EM, Green C, et al. Features of 20 133 UK patients in hospital with covid-19 using the ISARIC WHO Clinical Characterisation Protocol: prospective observational cohort study. BMJ. 2020;369:m1985. doi: 10.1136/bmj.m1985.

24. Berenguer J, Ryan P, Rodríguez-Baño J, Jarrín I, Carratalà J, Pachón J, Yllescas M, Arribas JR; COVID-19@Spain Study Group. Characteristics and predictors of death among 4,035 consecutively hospitalized patients with COVID-19 in Spain. Clin Microbiol Infect. 2020 Aug 4:S1198-743X(20)30431-6. doi: 10.1016/j.cmi.2020.07.024. Epub ahead of print.

25. Gharbharan A, Jordans C, Geurtsvankessel C et al. Convalescent plasma for COVID-19. A randomized clinical trial. medRxiv [Preprint] Jul 1, 2020 [cited 2020 Sep 25]. Available from doi: https://doi.org/10.1101/2020.07.01.20139857.

26. Agarwal A, Mukherjee A, Kumar G, et al. Convalescent plasma in the management of moderate COVID-19 in India: An open-label parallel-arm phase II multicentre randomized controlled trial (PLACID Trial). medRxiv [Preprint] Sep 3, 2020 [cited 2020 Sep 25] Available from doi: https://doi.org/10.1101/2020.09.03.20187252

27. Avendano-Sola C, Ramos-Martinez A, Munez-Rubio E et al. Convalescent Plasma for COVID-19: A multicenter, randomized clinical trial. medRxiv [Preprint] Aug 26, 2020 [cited 2020 Sep 25]. Available from doi: https://doi.org/10.1101/2020.08.26.20182444

28. Rasheed AM, Fatak DF, Hashim HA, et al. The therapeutic potential of convalescent plasma therapy on treating critically-ill COVID-19 patients residing in respiratory care units in hospitals in Baghdad, Iraq. Infez Med. 2020;28(3):357–366.

29. Xia X, Li K, Wu L, et al. Improved clinical symptoms and mortality among patients with severe or critical COVID-19 after convalescent plasma transfusion. Blood. 2020; 136(6): 755–759. doi:10.1182/blood.2020007079

30. Liu STH, Lin HM, Baine I, et al. Convalescent plasma treatment of severe COVID-19: a propensity score-matched control study. Nat Med. 2020. doi: 10.1038/s41591-020-1088-9. Epub ahead of print.

31. Zaki N, Alashwal H, Ibrahim S. Association of hypertension, diabetes, stroke, cancer, kidney disease, and high-cholesterol with COVID-19 disease severity and fatality: A systematic review. Diabetes Metab Syndr. 2020;14(5):1133–1142. doi:10.1016/j.dsx.2020.07.005

32. Jutzeler CR, Bourguignon L, Weis CV, et al. Comorbidities, clinical signs and symptoms, laboratory findings, imaging features, treatment strategies, and outcomes in adult and pediatric patients with COVID-19: A systematic review and meta-analysis [published online ahead of print, 2020 Aug 4]. Travel Med Infect Dis. 2020;37:101825. doi:10.1016/j.tmaid.2020.101825

33. Richardson S, Hirsch JS, Narasimhan M, et al; the Northwell COVID-19 Research Consortium.Presenting characteristics, comorbidities, and outcomes among 5700 patients hospitalized with COVID-19 in the New York City area. JAMA. 2020; 323(20):2052–2059. doi:10.1001/jama.2020.6775

34. Karagiannidis C, Mostert C, Hentschker C, et al. Case characteristics, resource use, and outcomes of 10?021 patients with COVID-19 admitted to 920 German hospitals: an observational study. Lancet Respir Med. 2020;8(9):853–862. doi:10.1016/S2213-2600(20)30316-7

